# Sarcopenia Assessment in Resource-Constrained Settings: Expert Agreement on Surrogate Measures

**DOI:** 10.64898/2026.09.06.26361926

**Authors:** Meghna Suresh Prabhu, Sucheta V Kolekar, Olivier Bruyere, Reshma A Merchant, Sanjay Kalra, Shweta Gore, Nimit Agarwal, Girish Nandakumar

**Author notes:** **corresponding author:** Email id, Contact details.

## Abstract

**Purpose:** Sarcopenia, the progressive loss of muscle mass and function with age, is diagnosed by assessing muscle strength, physical performance, and muscle mass. However, the tools required are often costly, inaccessible, or unsuitable for use in low-resource or remote settings. This study aimed to identify practical surrogate measures that are conceptually aligned with established sarcopenia diagnostic criteria and can be integrated into digital health platforms to enable remote screening and monitoring using an expert agreement process.

**Methods:** A narrative literature review was conducted to identify proxy measures for muscle strength (HGS), physical performance, and muscle mass. This was followed by two rounds of expert agreement. The Content Validity Index (CVI), defined as the number of experts in agreement divided by the total number of experts for each pragmatic proxy, was calculated after each round to assess the relevance and appropriateness of each measure. A pragmatic proxy with a CVI of 0.7 or higher was included.

**Results:** Eight proxies for HGS were identified across ten studies, with the five-times sit-to-stand test (5STS) being the most common. The 5STS has a higher agreement and was considered a relevant and appropriate surrogate by the expert panel for both muscle strength (CVI = 0.8 for both relevance and appropriateness) and physical performance (CVI = 1 for relevance and 0.8 for appropriateness), whereas the TUG could be a promising measure for physical performance (CVI = 1 for both relevance and appropriateness). Calf circumference (CC) offers a low-cost alternative for assessing muscle mass (CVI = 1 for relevance and 0.7 for appropriateness).

**Conclusion:** The 5STS, TUG, and CC were identified as relevant and appropriate pragmatic proxy measures for assessing sarcopenia, with potential for integration into digital screening tools in low-resource settings. Further clinical studies are required to establish their validity, reliability, and feasibility for remote assessment.

**Author Summary:** Sarcopenia is an age-related condition involving the loss of muscle strength, physical ability, and muscle mass. Identifying it usually requires specialised equipment and face-to-face assessment, which may not be available in remote or resource-limited settings. We therefore aimed to identify simple, practical measures that could support sarcopenia screening through digital health platforms.

We reviewed the existing literature to identify possible alternatives to conventional assessments of muscle strength, physical performance, and muscle mass. We then asked a panel of experts to evaluate these measures through a two-round agreement process based on their relevance and suitability. The experts identified the five-times sit-to-stand test, which measures how quickly a person can rise from a chair five times, as a practical indicator of muscle strength and physical performance. The Timed Up-and-Go test was considered a promising measure of physical performance, while calf circumference was identified as a simple, low-cost indicator of muscle mass.

Together, these measures could make sarcopenia screening more accessible and suitable for digital use in low-resource settings. Further clinical research is needed to determine whether they provide accurate, reliable, and feasible assessments when used remotely.

## INTRODUCTION

Sarcopenia is an age-related condition characterized by the progressive loss of muscle mass, strength, and function, leading to an increased risk of falls, frailty, and mortality (1). Recently, the life-course approach has gained recognition in understanding and managing sarcopenia. It emphasizes that muscle health is influenced by factors that operate throughout the lifespan, rather than only in later years (2). Consequently, early identification and timely preventive strategies are now considered crucial to delay or mitigate the onset and progression of sarcopenia. However, current estimates of sarcopenia prevalence vary widely (∼ 10% to 60%), primarily due to inadequate consistency in the operational definitions employed, the assessment tools utilized, and the demographic and clinical characteristics of the population studied (3–5). Moreover, available sarcopenia frameworks endorse the use of hand dynamometry, or tools such as bio-impedance analysis (BIA) and dual-energy x-ray absorptiometry (DEXA), which are largely equipment-dependent and therefore have limited use in remote or low-resource settings (2,6,7). Therefore, despite its clinical significance, early identification remains a challenge due to the high cost, time constraints, and expertise required for gold-standard diagnostic methods such as DEXA and magnetic resonance imaging (MRI) (8–10). Additionally, sarcopenia often goes underdiagnosed due to its gradual onset and overlap with normal aging, making large-scale community screening difficult, especially in low-resource settings, characterized by limited access to advanced imaging, calibrated dynamometry, trained specialists, and structured geriatric assessment infrastructure. (11).

Recent advancements in digital health technology offer potential solutions for community-level sarcopenia screening. Smartphone-based hand grip strength meters, wearable accelerometers, and AI-driven gait analysis are emerging as accessible alternatives to conventional assessments(12–14). These technologies generate a large volume of data, and machine learning models and AI-driven algorithms hold promise for refining these measures, enabling early detection and intervention (14,15). Standardized and reliable, pragmatic proxy measures are essential to ensure the accuracy and applicability of these novel screening approaches and to support the successful translation of smartphone- and wearable-based assessments into routine care in remote settings. These pragmatic proxy measures are not intended to replace the standard measures recommended by sarcopenia working groups; rather, they are proposed as pragmatic, functional measures for remote screening and monitoring in contexts where direct assessment of muscle strength or muscle mass is not feasible in order to enable a higher detection rate and improve accessibility across all geographical contexts.

In accordance with WHO’s strategies to advance health equity for all, integrating digital technology into healthcare offers a powerful means to expand access to diagnostic tests. With the increasing use of mobile devices and other technologies among older adults, the development of easy-to-use digital tests can help bridge the gap between undetected and detected cases of sarcopenia. Addressing this gap is critical to advancing digital screening and remote monitoring for older adults in low-resource settings. Therefore, the objective of this study was to identify and establish agreement on functional pragmatic proxy measures of sarcopenia that are conceptually aligned with established diagnostic criteria and can be integrated into digital platforms to support remote, scalable, and low-cost screening in aging populations.

## METHODS

The study followed a sequential exploratory design, and the protocol was approved by the Institutional Ethics Committee, Kasturba Medical College and Kasturba Hospital, Manipal Academy of Higher Education, Manipal, Karnataka, India [IEC1: 179], and was prospectively registered on the Clinical Trial Registry-India (CTRI) platform on 18^th^ October 2024. (CTRI/2024/10/075509). This study was done in two steps: a) identifying pragmatic proxy measures through a narrative literature review, and b) expert agreement process to determine relevance and appropriateness for digital integration.

### a. Identifying pragmatic proxy measures

**Figure 1:**
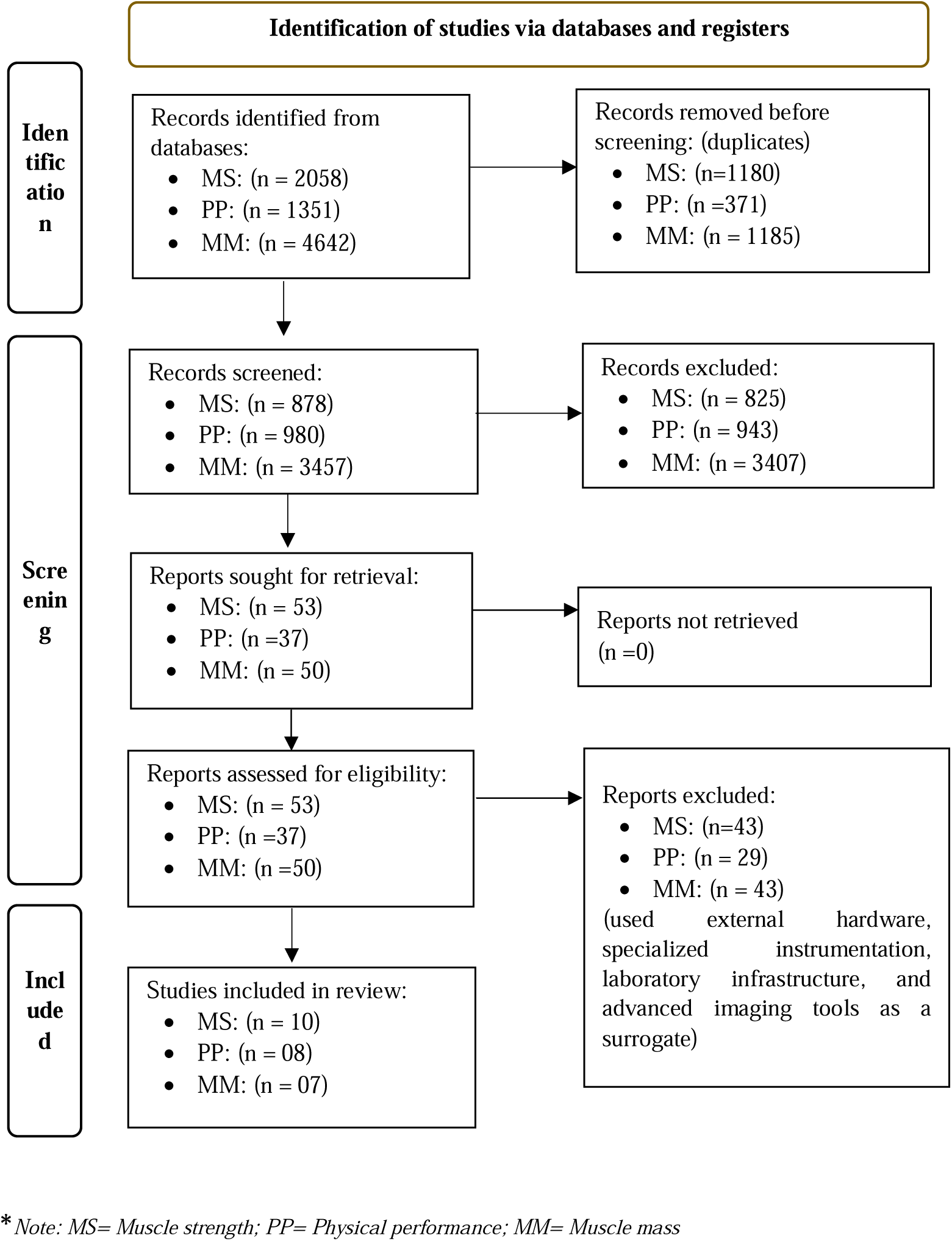
PRISMA flow diagram.

A narrative literature review was undertaken to identify functional pragmatic proxy measures that are conceptually aligned with established sarcopenia diagnostic criteria (muscle strength, physical performance, and muscle mass), and potentially adaptable for digital integration. This study utilized the Asian Working Group for Sarcopenia 2019 (AWGS 2019) sarcopenia diagnostic criteria as the reference standard to identify functional pragmatic proxy measures contextualized for screening and/or risk identification, rather than as diagnostic equivalents, to understand their potential clinical implications in remote or resource-limited settings. The search was conducted using pre-validated keywords across five databases, namely, CINAHL, PubMed, Embase, Scopus, and Web of Science. The search strategy was peer-reviewed for validation using the Peer Review of Electronic Search Strategies (PRESS) checklist, developed by Health Technology Review (2015), and independently assessed by two reviewers. The keywords, search strategy, and data extraction process have been detailed in Online Resource 1.

The selection criteria were defined as follows: Studies were included if they met the following criteria: (a) involved older adults aged ≥ 60 years; (b) were published in the English language; (c) comprised original peer-reviewed research, including cross-sectional, cohort, case–control, diagnostic accuracy, validation, or interventional study designs; (d) reported pragmatic proxy measures corresponding to at least one established sarcopenia domain (muscle strength, physical performance, or muscle mass) with comparison against recognized reference standards (e.g., handgrip strength, Short Physical Performance Battery (SPPB), Dual-energy X-ray Absorptiometry (DXA), Bio-electrical Impedance Analyzer (BIA), Computed Tomography (CT), or Magnetic Resonance Imaging(MRI)); and (e) evaluated measures with potential applicability in a resource constrained setting, for remote assessment, including self-administration or compatibility with smartphone-based platforms. Studies were excluded if they: (a) consisted of editorials, conference abstracts, reviews, or case reports without original empirical data; and (b) evaluated pragmatic proxy measures requiring external hardware, specialised instrumentation, laboratory-based infrastructure, or advanced imaging modalities that are not feasible for remote assessment or digital integration through smartphone-based or wearable technologies.

### b. Expert agreement process

A panel of experts was identified based on their representation in sarcopenia consensus statements and international working groups, as well as their affiliations with geriatric organizations, including the British Geriatric Society (BGS), the American Geriatrics Society (AGS), and the National Institute on Aging (NIA). For this study, expertise was defined as scholarly publications on sarcopenia, geriatric medicine, rehabilitation, or muscle health; participation in consensus statements, guideline panels, or recognized geriatric organizations; and at least 10 years of professional experience in geriatrics or related fields. Additionally, an h-index ≥ 4, verified via a Scopus author profile, was used as an objective indicator of research impact. An invitation email was sent to prospective experts, which included an overview of the research objectives, the rationale for this study, a description of the measure identification process, and an outline of their expected role. It also detailed the structure of the agreement rounds and the rating score to be used. Experts who consented to participate were subsequently sent survey links via email.

The functional pragmatic proxies identified in Step A were compiled and presented to the expert panel through three separate Microsoft Forms, covering muscle strength, muscle mass, and physical performance measures relevant to sarcopenia assessment. The experts were required to evaluate each listed pragmatic proxy using a four-point Likert scale, assessing the measure in two dimensions: the relevance of the item to its respective domain (muscle strength, muscle mass, or physical performance) and its appropriateness for digital integration. The four-point Likert scale was defined as follows for both domains: Strongly Agree, Agree, Disagree, Strongly Disagree. The neutral option was deliberately excluded to avoid ambiguity and encourage respondents to provide more definitive opinions. Each survey required approximately 10-15 minutes to complete. The forms are attached as Online Resource 2.

Expert agreement was established through a two-round iterative process. In Round 1, experts evaluated the potential pragmatic proxy measures identified through the narrative literature review and could recommend additional measures using open-ended responses. Participants received a summary of the review methodology and detailed instructions for rating each measure. Once all responses had been received, a Content Validity Index (CVI) was calculated for each measure by dividing the number of experts who endorsed it by the total number of experts who evaluated it. Measures with a CVI of ≥0.70 were retained for Round 2. Any additional measure frequently recommended by the expert panel was also considered for inclusion. To maximise participation, two reminder emails were sent at seven-day intervals during Round 1.

In Round 2, experts reassessed the refined set of measures selected based on the Round 1 CVI scores and qualitative recommendations. A summary of the first-round results and panel feedback was provided to inform their reassessment. Experts then indicated their final approval of each measure and offered any further recommendations. To account for agreement occurring by chance, the modified kappa statistic (k*) was calculated for each measure in both the relevance and appropriateness domains. Modified kappa values were interpreted as excellent (>0.74), good (0.60–0.74), fair (0.40–0.59), or poor (<0.40). By integrating CVI and chance-corrected agreement with iterative qualitative feedback, this process enabled the selection of feasible, contextually appropriate pragmatic proxy measures for inclusion in the digital tool.

## RESULTS

### a. Identifying pragmatic proxy measures

A total of 8,051 records were identified, 2,058 for muscle strength, 1,351 for physical performance, and 4,642 for muscle mass. After initial screening, 140 records were retrieved for full-text screening (53 for muscle strength, 37 for physical performance, and 50 for muscle mass). Ultimately, 25 studies were identified (10 for muscle strength, 08 for physical performance, and 07 for muscle mass), which reported different pragmatic proxy measures based on the inclusion criteria.

#### i) Muscle strength

Ten studies reported eight different measures used as proxies for handgrip strength to assess muscle strength. The measures reported are as follows: Five times sit-to-stand test (5STS) (16,17), 30-second chair stand test (30-s CST) (18,19), 10 times sit-to-stand test (20,21), regression equation (21), blink rate (22), VibPress (23), calf-raise senior test (24), and the Sit-to-stand app (25,26). The details of the studies have been reported in Online Resource 3.

#### ii) Physical Performance

Eight studies reported the use of five different measures to assess physical performance. The measures are as follows: Five times sit-to-stand test (27–30), Functional Fitness test (31), gait speed (32), anthropometric measures (33), and eSPPB software (34). The details of the studies have been reported in Online Resource 4.

#### iii) Muscle Mass

Seven studies were identified, with calf circumference (CC) being the most reported measure. Most of these studies reported the cut-off for calf circumference between 32-36 cm, with a few outliers being reported at 29cm and 39cm (35–41). The details of the studies have been reported in Online Resource 5.

### b. Expert agreement process

The pragmatic proxy measures identified from the narrative literature review were listed in Microsoft Forms in descending order based on their frequency of occurrence across the included articles. Thirty-four experts (n = 34) from various geographical regions were contacted via email. Nine experts (n=9; 26.4%) had agreed to participate in the agreement process, of whom eight responded for both rounds of the expert agreement. The distribution of experts was as follows: Europe (Belgium (n=2) and Spain (n=1)), USA (n=2), Australia (n=1), and Asia (n=2), (Singapore (n=1), India (n=1)). The details of the experts have been attached as Online Resource 6. The identified measures were listed in the form, along with their DOIs, for the experts’ reference. The CVI was calculated at the end of both Round 1 and Round 2.

**Table 1:** List of measures for expert agreement:

| Muscle strength | Physical performance | Muscle mass |
| --- | --- | --- |
| • Five times sit-to-stand test (5STS)<br>• 30-second chair stand test<br>• 10 times sit-to-stand test (10STS)<br>• Regression equation<br>• Blink rate<br>• VibPress<br>• Calf-raise senior test (CRS test)<br>• Sit-to-stand app | • Five times sit-to-stand test<br>• Functional Fitness test<br>• Gait speed<br>• Anthropometric measures<br>• eSPPB software | • Calf circumference (CC) |

**Table 2:**
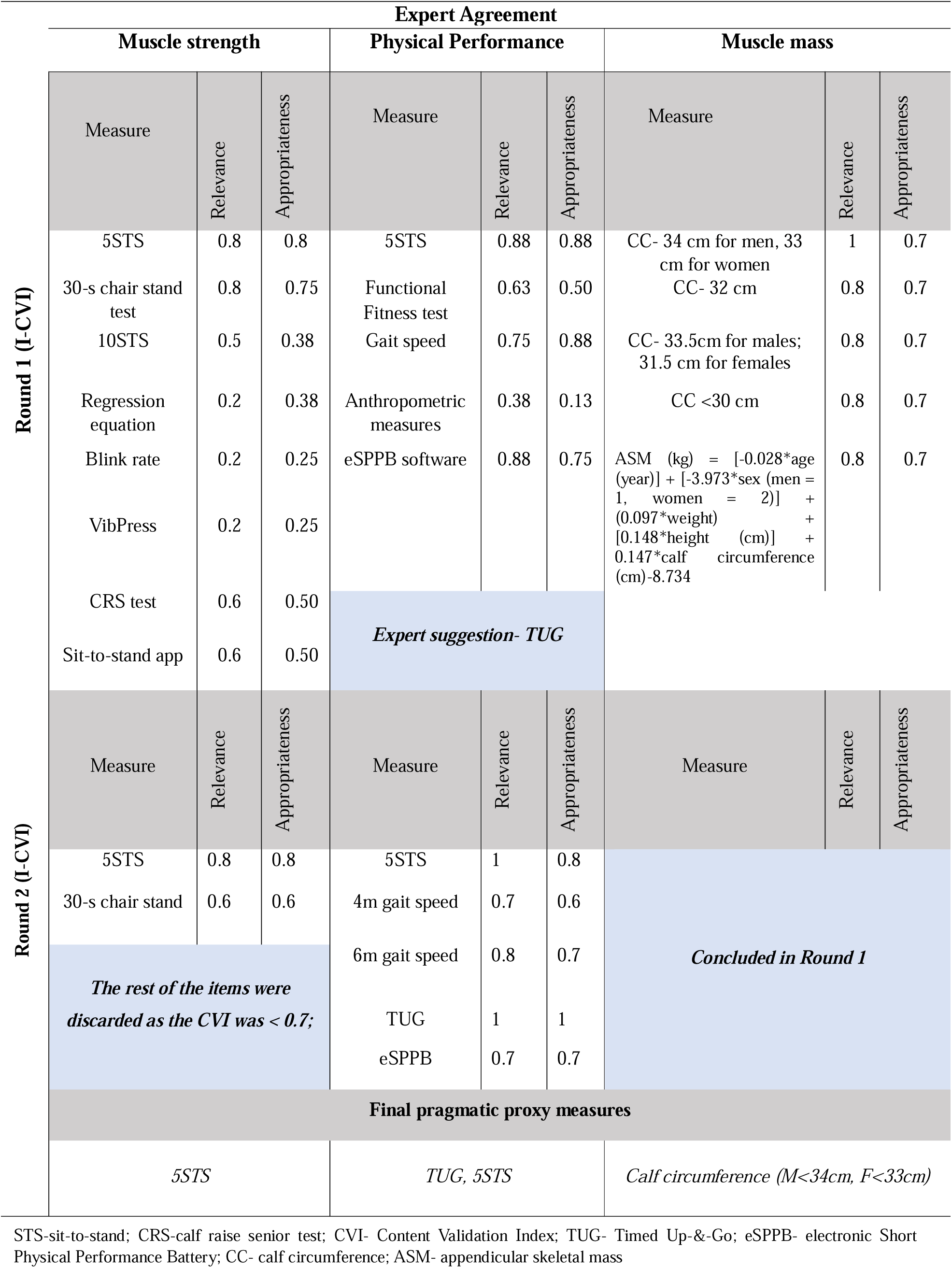
Flowchart for expert agreement.

### Recommended measures for muscle strength

In round 1, the form included eight (n=8) listed measures and one section for the experts’ suggestions. Six measures (n = 6) did not exceed the preset value of CVI > 0.7 for appropriateness and relevance, namely, 10 times sit-to-stand, regression equation, blink rate, VibPress, calf-raise senior test, and sit-to-sand app, and were therefore excluded after the first round. Two measures (n = 2), namely, the five-times sit-to-stand test (5STS) and the 30-second chair stand test, had a CVI value of ≥ 0.7 and were carried forward to round 2. Four experts did not provide additional suggestions. The other four provided the following suggestions: “biomarkers”, “subjective handgrip strength when handshaking”, “Timed Up-and-Go test (TUG) test”, and “Handgrip”. In round 2, the 5STS test had a CVI of 0.8 for both appropriateness and relevance, while the 30-second chair stand test had a CVI of 0.6 for both domains. Although both items were equally favoured by the eight experts, the higher CVI value of 5STS indicates a stronger consensus as a proxy measure of muscle strength.

In round 1, the modified kappa statistic (k*) indicated excellent agreement for both the 5STS and the 30s-CST in the relevance domain (k* = 0.87). The remaining measures showed fair-to-poor agreement (k* = 0.52–0.20). In the appropriateness domain, agreement was excellent for the 5STS (k* = 0.87), whereas the other measures showed agreement ranging from good to poor (k* = 0.72–0.15). In round 2, agreement remained excellent for the 5STS (k* = 0.87) but was fair for the 30s-CST (k* = 0.52) across both domains.

### Recommended measure for physical performance

In round 1, the form included five (n=5) listed measures and one section for the experts’ opinions or suggestions. Two measures (n=2), i.e., the functional fitness test and anthropometric measures, had CVIs <0.7 in both domains and were therefore excluded after the first round. Three measures (n = 3), namely, the 5STS, the gait speed test, and the electronic version of the SPPB (eSPPB), had a CVI of ≥0.7 and were carried forward to the second round. In the section for the expert’s suggestions, four authors did not provide additional suggestions, while the remaining four suggested the TUG and further differentiation for 4m and 6m gait speed. TUG did not emerge from our initial pragmatic proxy-focused search strategy tailored to the AWGS 2019 guidelines, but it is a well-documented measure of physical performance, is endorsed in the European frameworks and clinical studies, (42,43) and has accordingly been carried forward for round 2.

In round 2, the measures included were: 5STS, gait speed- 4m and 6m, and eSPPB. The 5STS test had a CVI of 1 in the relevance domain and 0.8 in the appropriateness domain, while the TUG test had a CVI of 1 in both domains. The 6m gait speed test had a CVI of 0.8 in the relevance domain and 0.7 in the appropriateness domain. The other two measures, i.e., the 4m gait speed test and the eSPPB test, had a CVI of <0.8 in both domains. In the ranking preferences, the 5STS, TUG, and eSPPB received the most favourable ranking. However, because the CVIs for the 5STS and TUG are the highest, both measures can be considered pragmatic proxy markers of SPPB for physical performance in older adults.

In round 1, the modified kappa statistic indicated excellent agreement for 5STS and eSPPB (k* = 0.87), good agreement for gait speed (k* = 0.72), fair agreement for functional fitness (k* = 0.53), and poor agreement for anthropometric measures (k* = 0.21). In round 2, excellent agreement was observed for 5STS and gait speed (k* = 0.87), while eSPPB demonstrated good agreement (k* = 0.72).

### Recommended measures for muscle mass

For muscle mass, we received responses from only seven (n=7) experts. One expert did not complete the form for pragmatic proxy measures of muscle mass. There was no response, even after the second reminder. In round 1, experts evaluated four listed measures (n = 4). Calf circumference was the most frequently recommended additional measure, with proposed cut-off values ranging from 32 to 36 cm. The most common cut-off of 34 cm received a higher consensus. Calf circumference with a cut-off value of <34 cm in men and <33 cm in women had the highest CVI of 1 for relevance and 0.7 for appropriateness. The other items also had a CVI of > 0.7; however, since the cut-off value of < 34cm in men and < 33cm in women had a clear agreement between all seven experts, it was decided to conclude the validation process at round 1 itself. Five experts (n = 5) did not provide any additional recommendations for muscle mass, while one expert (n = 1) suggested using body indices such as the Waist-to-Calf Ratio (WCR), CCI (Calf Central Index), and BCI (Body Calf Index). The modified kappa statistic indicated excellent agreement regarding relevance for all four items. Calf circumference (<34 cm in men and <33 cm in women) achieved perfect agreement beyond chance (k* = 1.00), while Items 2, 3, and 4 each demonstrated excellent agreement (k* = 0.85). Regarding appropriateness, all four items demonstrated good agreement (k* = 0.64).

Through this expert agreement process, we identified key assessment tools that could serve as potential pragmatic proxy measures for detecting sarcopenia. The 5STS was proposed as a suitable indicator of muscle strength, reflecting functional muscle use in everyday activities. Both the 5STS and the Timed Up-and-Go (TUG) test were similarly endorsed as reliable indicators of physical performance, given their ability to assess balance, gait speed, and functional mobility. Lastly, the calf circumference measurement was purported to be a simple and non-invasive method for estimating muscle mass.

## DISCUSSION

In recent years, the pursuit of alternative approaches for assessing sarcopenia has intensified, particularly in response to the limitations of conventional methods that depend on expensive equipment and specialized personnel, which are often unavailable in resource-constrained settings (44,45). The AWGS 2025 consensus emphasizes the importance of focusing on evaluating muscle health in midlife, thereby promoting early intervention. It also underscores the need for regular assessments, at both the clinical and community levels(2). Advances in technology, particularly Artificial Intelligence (AI), Internet of Things (IoT), and sensor-based tools, offer promising opportunities to improve accessibility, enable remote monitoring, and support early detection, especially for older adults with mobility challenges(46–48). This study contributes to the field by identifying evidence-based, feasible, and potentially integrable pragmatic proxy measures for community and home settings, which can be implemented on mHealth platforms in resource-constrained environments.

Through a narrative literature review and expert agreement process, multiple candidate measures of muscle strength, physical performance, and muscle mass were evaluated. Although the literature search was not specifically designed to assess digital feasibility, the expert panel additionally considered the potential suitability of each measure for integration into digital platforms. Several device-dependent approaches were identified in the literature but were excluded because they required specialised equipment or trained personnel, limiting their suitability for remote or resource-constrained assessment. These included measures of lip strength, modified sphygmomanometer-based strength testing, the Gripwise digital dynamometer, oral-function assessments, and instrumented knee-extensor strength testing. (49–52). One study also explored the Borg CR10 scale to estimate grip force during hand-tool use. Although these approaches may provide alternative estimates of muscle strength, their dependence on additional equipment or specialised administration reduces their practicality for scalable, low-resource digital screening.

Recent consensus frameworks, including the Global Leadership Initiative in Sarcopenia (GLIS) and AWGS 2025, place greater emphasis on muscle strength and muscle mass, with physical performance viewed primarily as an indicator of functional impact rather than a defining component of sarcopenia. (53; 6). Nevertheless, in low-resource and remote settings, functional decline may be the first observable manifestation prompting further assessment. Physical performance measures were therefore retained in this study as pragmatic screening indicators rather than diagnostic surrogates.

The expert consensus identified the 5STS as a pragmatic proxy measure of muscle strength. This finding is consistent with the EWGSOP2 recommendations, which recognise the 5STS as an alternative when handgrip-strength assessment is not feasible (7). Although the 5STS primarily assesses lower-limb function rather than muscle strength in isolation, it provides an indirect indication of overall muscular performance and reflects abilities essential to daily living, including rising from a chair, maintaining balance, and performing routine mobility tasks. Handgrip strength remains a widely used objective measure; however, it requires specialised equipment, limiting its feasibility for integration into a digital health tool, particularly in low-resource settings. The 5STS offers a simple and equipment-free alternative, although its results should be interpreted cautiously in individuals with substantial pain or balance and mobility impairments.

Regarding physical performance, 5STS and TUG received the strongest expert endorsement. TUG did not emerge from the initial literature search, which was based on the AWGS 2019 framework, but was subsequently proposed by the expert panel and retained for Round 2. Although TUG is widely used to assess functional mobility and is associated with outcomes such as falls, frailty, and functional decline, its role specifically as a screening measure for sarcopenia remains less well established. Further studies are therefore needed to evaluate its diagnostic accuracy, clinical utility, and feasibility for remote or digital assessment.

For muscle mass, calf circumference was the most commonly identified anthropometric proxy and received the strongest expert endorsement. Previous studies have reported associations between calf circumference and appendicular skeletal muscle mass, supporting its potential utility as a simple screening measure. Other approaches included predictive equations combining anthropometric variables with handgrip strength, as well as temporal muscle thickness (54); however, these approaches were considered less suitable for remote or resource-constrained assessment because of their additional measurement requirements. Overall, the expert panel supported calf circumference as a pragmatic, non-invasive measure for large-scale screening where direct assessment of muscle mass is not feasible.

Overall, this study identified a concise set of pragmatic proxy measures-5STS, TUG, and CC for potential use in resource-constrained settings. Their simplicity and minimal equipment requirements may make them suitable candidates for future remote or digitally supported screening. However, further clinical validation is required to establish their reliability, diagnostic accuracy, safety, and feasibility when used outside conventional face-to-face assessment. Given the challenges of implementing standard sarcopenia assessments in many low-resource settings, these measures may offer a practical approach to identifying individuals who warrant further evaluation.

## SIGNIFICANCE

The growing number of older adults worldwide highlights the need for accessible approaches to identifying sarcopenia, particularly in resource-constrained settings where standard assessment tools may not be readily available. This study identifies pragmatic proxy measures that may support scalable screening when conventional assessment of muscle strength, physical performance, and muscle mass is difficult to implement. Their simplicity and limited equipment requirements also make them potential candidates for future adaptation to digital or remotely supported assessment. Further validation is required to establish their reliability, diagnostic accuracy, and feasibility when implemented through digital platforms.

## LIMITATIONS

First, the pragmatic proxy measures were identified solely from published articles in five electronic databases, potentially excluding other relevant sources. Second, the expert panel was relatively small, and the low response rate may have introduced non-response bias and limited the representativeness of the findings. Although several panel members were based in high-income countries, some had clinical, research, or implementation experience in resource-constrained settings. Nevertheless, representation from experts currently practising in low- and middle-income countries was limited and was not proportionally stratified, which may have reduced the diversity of perspectives specific to these settings. Variation in sarcopenia definitions and reference standards across the included studies also limited direct comparability between candidate measures. Finally, although experts were provided with descriptions and references for each measure, familiarity with individual measures was not formally assessed and may have influenced their ratings.

## FUTURE RECOMMENDATIONS

The growing demand for scalable, accessible assessment tools for sarcopenia in primary care and community settings makes this study a valuable contribution, as it identifies measures that are not only cost-effective but also better reflect functional status and real-world outcomes. Future studies should prospectively evaluate the identified proxy measures in primary care, community, and home settings. This should include assessment of reliability, diagnostic accuracy, feasibility, safety, and acceptability, particularly when the measures are self-administered or remotely supervised. Further work is also required to determine whether these measures can be reliably adapted to digital platforms, including video- or sensor-assisted assessment, and whether performance differs from conventional face-to-face administration. Validation across diverse populations and resource settings will be important before their use in large-scale sarcopenia screening can be recommended.

## CONCLUSION

This study identified a range of candidate proxy measures for assessing muscle strength, physical performance, and muscle mass in older adults. Expert agreement supported 5STS as a practical proxy for muscle strength, TUG as a promising measure of physical performance, and CC as a pragmatic proxy for muscle mass. Together, these measures may offer a simple, low-resource approach to sarcopenia screening in settings where conventional assessment tools are not readily available. Further clinical and digital validation is required before their use in remote or large-scale screening can be recommended.

## Supporting information

Online resource 1

Online resource 2

Online resource 3

Online resource 4

Online resource 5

Online resource 6

## Data Availability

All relevant data are within the manuscript and its Supporting Information files.

## ACKNOWLEDGEMENT

The authors wish to acknowledge the use of Grammarly to improve written English and Grammar, Trinka AI for paraphrasing, and Mendeley for citation purposes.

## ONLINE RESOURCE

Online Resource 1- Keywords and search strategy

Online Resource 2- Microsoft Forms

Online Resource 3- Measures identified for muscle strength

Online Resource 4- Measures identified for physical performance

Online Resource 5- Measures identified for muscle mass

Online Resource 6- Details of experts

