## Supplementary material for "Sarcopenia Assessment in Resource-Constrained Settings: Expert Agreement on Surrogate Measures": Online resource 1

**Title: Identification and Expert Agreement on Surrogate Measures for Sarcopenia in Older Adults in Low-Resource Settings**

**Journal: European Geriatric Medicine**

**Authors: Ms. Meghna Suresh Prabhu, Dr. Sucheta V Kolekar, Olivier Bruyere, Reshma A Merchant, Sanjay Kalra, Shweta Gore, Nimit Agarwal, Dr Girish N***

***Dr. Girish N (PhD) (corresponding author),**

Additional Professor,

Department of Physiotherapy, Manipal College of Health Professions,

Manipal Academy of Higher Education,

Karnataka, India.

ORCID: 0000-0003-2181-5332

Contact details: +91 9886782114

**Online Resource 1- Keywords & search strategy.**

i. *Keywords:*

Included various combinations of terms such as 'muscle strength,' 'hand grip strength,' 'grip strength,' 'muscle mass,' 'DEXA,' 'BIA,' 'physical performance,' 'short physical performance battery,' 'SPPB,' 'gait speed,' 'chair stand test,' 'surrogate measure,' 'proxy measure,' and 'alternate measure,' combined using 'AND' and 'OR' Boolean operators.

ii. *Data extraction:*

The search was conducted separately for each measure, and the records were imported into Rayyan as three separate reviews to make the screening process easier (#1054743, #1054745, #1066857). After de-duplication, two reviewers independently screened titles/abstracts and full texts, with disagreements resolved by a third reviewer. A data extraction chart was developed by two reviewers, validated by the remaining reviewers, and refined accordingly. Data were extracted on study characteristics, sarcopenia diagnosis and criteria, surrogate measures and assessment procedures, anthropometric data, statistical analyses, diagnostic accuracy, and key findings.

iii. *Search strategy:*

Search strategy for surrogate measures-

1. Muscle strength

a. (surrogate measures OR proxy measures OR alternate measures) AND (hand grip strength)

b. (surrogate measures OR proxy measures OR alternate measures) AND (grip strength)

c. (surrogate measures OR proxy measures OR alternate measures) AND (hand strength)

2. Physical performance

a. (surrogate measures OR proxy measures OR alternate measures) AND (gait speed)

b. (surrogate measures OR proxy measures OR alternate measures) AND (chair stand test OR sit-to-stand test or 5 times sit-to-stand test)

c. (surrogate measures OR proxy measures OR alternate measures) AND (SPPB OR Short Physical Performance Battery)

3. Body composition

a. (surrogate measures OR proxy measures OR alternate measures) AND (body composition)

b. (surrogate measures OR proxy measures OR alternate measures) AND (lean muscle mass)

c. (surrogate measures OR proxy measures OR alternate measures) AND (appendicular skeletal muscle mass)

d. (surrogate measures OR proxy measures OR alternate measures) AND (skeletal muscle mass)

*Results with search string as used*-

**MUSCLE STRENGTH**

| Sr. No. | Concept | Sub-heading | Database | Search strategy with filter | Results |
| --- | --- | --- | --- | --- | --- |
| 1 | Muscle strength | 1a | CINAHL Ultimate | ((surrogate measures OR proxy measures OR alternate measures)) AND (hand grip strength) | 12 |
|  |  |  | Embase | ((('surrogate'/exp OR surrogate) AND measures OR 'proxy'/exp OR proxy) AND measures OR alternate) AND measures AND ('hand'/exp OR hand) AND grip AND ('strength'/exp OR strength) | 67 |
|  |  |  | PubMed | ((surrogate measures OR proxy measures OR alternate measures)) AND ((hand grip strength)) AND ((english[Filter]) AND (aged[Filter] OR middleaged[Filter] OR 80andover[Filter])) | 378 |
|  |  |  | Scopus | ( surrogate measures OR proxy measures OR alternate measures ) AND ( hand grip strength ) | 76 |
|  |  |  | Web Of Science | (ALL=((surrogate measures OR proxy measures OR alternate measures))) AND ALL=((hand grip strength)) | 78 |
|  | Muscle strength | 1b | CINAHL Ultimate | ((surrogate measures OR proxy measures OR alternate measures)) AND (grip strength) | 17 |
|  |  |  | Embase | ('surrogate measures' OR 'proxy measures' OR 'alternate measures') AND ('grip strength'/exp OR 'grip strength') | 8 |
|  |  |  | PubMed | (((surrogate measures OR proxy measures OR alternate measures))) AND ((grip strength)) AND ((english[Filter]) AND (middleaged[Filter] OR aged[Filter] OR 80andover[Filter])) | 446 |
|  |  |  | Scopus | ( "surrogate measures" OR "proxy measures" OR "alternate measures" ) AND ( "grip strength" ) | 344 |
|  |  |  | Web Of Science | (ALL=((surrogate measures OR proxy measures OR alternate measures))) AND ALL=((grip strength)) | 180 |
|  | Muscle strength | 1c | CINAHL | ( ('surrogate measures' OR 'proxy measures' OR 'alternate measures') ) AND ('hand strength') | 12 |
|  |  |  | Embase | ('surrogate measures' OR 'proxy measures' OR 'alternate measures') AND ('hand strength'/exp OR 'hand strength') | 10 |
|  |  |  | PubMed | ((surrogate measures OR proxy measures OR alternate measures)) AND ((hand strength)) AND ((english[Filter]) AND (middleaged[Filter] OR aged[Filter] OR 80andover[Filter])) | 378 |
|  |  |  | Scopus | ( "surrogate measures" OR "proxy measures" OR "alternate measures" ) AND ( "hand strength" ) | 56 |
|  |  |  | Web of Science | ALL=((surrogate measures OR proxy measures OR alternate measures) AND (hand strength)) and Rehabilitation or Geriatrics Gerontology or Gerontology (Web of Science Categories) | 38 |

**PHYSICAL PERFORMANCE**

| Sr. No. | Context | Sub-heading | Database | Search strategy with filter | Results |
| --- | --- | --- | --- | --- | --- |
| 2 | Physical performance | 2a | CINAHL | ((surrogate measures OR proxy measures OR alternate measures)) AND (gait speed) | 13 |
|  |  |  | Embase | ('surrogate measures' OR 'proxy measures' OR 'alternate measures') AND ('gait speed'/exp OR 'gait speed') | 18 |
|  |  |  | Pubmed | ((surrogate measures OR proxy measures OR alternate measures)) AND ((gait speed)) AND ((english[Filter]) AND (middleaged[Filter] OR aged[Filter] OR 80andover[Filter])) | 307 |
|  |  |  | Scopus | ( "surrogate measures" OR "proxy measures" OR "alternate measures" ) AND ( "gait speed" ) | 344 |
|  |  |  | Web of Science | ALL=((surrogate measures OR proxy measures OR alternate measures) AND (gait speed)) | 214 |
|  | Physical performance | 2b | CINAHL | ( (surrogate measures OR proxy measures OR alternate measures) ) AND ( (chair stand test OR sit-to-stand test or 5 times sit-to-stand test) ) | 4 |
|  |  |  | Embase | ((('surrogate'/exp OR surrogate) AND measures OR 'proxy'/exp OR proxy) AND measures OR alternate) AND measures AND ((('chair'/exp OR chair) AND ('stand'/exp OR stand) AND ('test'/exp OR test) OR 'sit to stand'/exp OR 'sit to stand') AND ('test'/exp OR test) OR 5) AND times AND ('sit to stand'/exp OR 'sit to stand') AND ('test'/exp OR test) | 6 |
|  |  |  | Pubmed | (( surrogate AND measures OR proxy AND measures OR alternate AND measures )) AND ((chair AND stand AND test OR sit-to-stand AND test OR 5 times AND sit-to-stand AND test )) AND ((english[Filter]) AND (middleaged[Filter] OR aged[Filter] OR 80andover[Filter])) | 56 |
|  |  |  | Scopus | ( surrogate AND measures OR proxy AND measures OR alternate AND measures ) AND ( chair AND stand AND test OR sit-to-stand AND test OR 5 times AND sit-to-stand AND test ) | 36 |
|  |  |  | Web of Science | (ALL=((surrogate measures OR proxy measures OR alternate measures))) AND ALL=((chair stand test OR sit-to-stand test or 5 times sit-to-stand test)) | 68 |
|  |  | 2c | CINAHL | ( (surrogate measures OR proxy measures OR alternate measures ) AND ( (SPPB OR Short Physical Performance Battery) ) | 6 |
|  |  |  | EMBASE | ((('surrogate'/exp OR surrogate) AND measures OR 'proxy'/exp OR proxy) AND measures OR alternate) AND measures AND ((('chair'/exp OR chair) AND ('stand'/exp OR stand) AND ('test'/exp OR test) OR 'sit to stand'/exp OR 'sit to stand') AND ('test'/exp OR test) OR 5) AND times AND ('sit to stand'/exp OR 'sit to stand') AND ('test'/exp OR test) | 40 |
|  |  |  | Pubmed | ((surrogate measures OR proxy measures OR alternate measures)) AND ((SPPB OR Short Physical Performance Battery)) AND ((english[Filter]) AND (middleaged[Filter] OR aged[Filter] OR 80andover[Filter])) | 47 |
|  |  |  | Scopus | ( "surrogate measures" OR "proxy measures" OR "alternate measures" ) AND ( sppb OR "Short Physical Performance Battery" ) | 180 |
|  |  |  | Web of Science | (ALL=((surrogate measures OR proxy measures OR alternate measures) )) AND ALL=((SPPB OR Short Physical Performance Battery)) | 27 |

**3. MUSCLE MASS**

| Sr. No. | Context | Sub-heading | Database | Search strategy with filter | Results |
| --- | --- | --- | --- | --- | --- |
| 3 | Muscle mass | 3a | CINAHL | ( (surrogate measures OR proxy measures OR alternate measures) ) AND (body composition) | 51 |
|  |  |  | EMBASE | ('surrogate measures' OR 'proxy measures' OR 'alternate measures') AND ('body composition'/exp OR 'body composition') | 110 |
|  |  |  | Pubmed | ((surrogate measures OR proxy measures OR alternate measures)) AND ((body composition)) AND ((english[Filter]) AND (middleaged[Filter] OR aged[Filter] OR 80andover[Filter])) | 716 |
|  |  |  | Scopus | ( "surrogate measures" OR "proxy measures" OR "alternate measures" ) AND ( "body composition" ) | 1434 |
|  |  |  | Web of Science | **(ALL=((surrogate measures OR proxy measures OR alternate measures))) AND ALL=((body composition))** and **Sport Sciences** or **Geriatrics Gerontology** or **Rehabilitation** or **Gerontology** or **Nursing** (Web of Science Categories) | 88 |
|  |  | 3b | CINAHL | ( (surrogate measures OR proxy measures OR alternate measures) ) AND (lean muscle mass) | 3 |
|  |  |  | Embase | ((('surrogate'/exp OR surrogate) AND measures OR 'proxy'/exp OR proxy) AND measures OR alternate) AND measures AND lean AND ('muscle'/exp OR muscle) AND ('mass'/exp OR mass) | 70 |
|  |  |  | Pubmed | ((surrogate measures OR proxy measures OR alternate measures)) AND ((lean muscle mass)) AND ((english[Filter]) AND (middleaged[Filter] OR aged[Filter] OR 80andover[Filter])) | 99 |
|  |  |  | Scopus | ("surrogate measures" OR "proxy measures" OR "alternate measures") AND ("lean muscle mass") | 1348 |
|  |  |  | WoS | (ALL=((surrogate measures OR proxy measures OR alternate measures))) AND ALL=((lean muscle mass)) | 139 |
|  |  | 3c | CINAHL | ( (surrogate measures OR proxy measures OR alternate measures) ) AND (appendicular skeletal muscle mass) | 1 |
|  |  |  | Embase | ('surrogate measures' OR (('surrogate'/exp OR surrogate) AND measures) OR 'proxy measures' OR (('proxy'/exp OR proxy) AND measures) OR 'alternate measures' OR (alternate AND measures)) AND ('appendicular skeletal muscle mass'/exp OR 'appendicular skeletal muscle mass' OR (appendicular AND skeletal AND ('muscle'/exp OR muscle) AND ('mass'/exp OR mass))) | 17 |
|  |  |  | Pubmed | ((surrogate measures OR proxy measures OR alternate measures)) AND ((appendicular skeletal muscle mass)) AND ((english[Filter]) AND (middleaged[Filter] OR aged[Filter] OR 80andover[Filter])) | 48 |
|  |  |  | Scopus | ( "surrogate measures" OR "proxy measures" OR "alternate measures" ) AND ( "appendicular skeletal muscle mass" ) | 32 |
|  |  |  | WoS | **(ALL=((surrogate measures OR proxy measures OR alternate measures))) AND ALL=((appendicular skeletal muscle mass)** | 34 |
|  |  | 3d | CINAHL | ( (surrogate measures OR proxy measures OR alternate measures) ) AND (skeletal muscle mass) | 4 |
|  |  |  | Embase | **('surrogate measures' OR (('surrogate'/exp OR surrogate) AND measures) OR 'proxy measures' OR (('proxy'/exp OR proxy) AND measures) OR 'alternate measures' OR (alternate AND measures)) AND ('skeletal muscle mass'/exp OR 'skeletal muscle mass' OR (skeletal AND ('muscle'/exp OR muscle) AND ('mass'/exp OR mass)))** | 106 |
|  |  |  | Pubmed | **((surrogate measures OR proxy measures OR alternate measures)) AND ((skeletal muscle mass)) AND ((english[Filter]) AND (middleaged[Filter] OR aged[Filter] OR 80andover[Filter]))** | 253 |
|  |  |  | Scopus | **( "surrogate measures" OR "proxy measures" OR "alternate measures" ) AND ( "skeletal muscle mass")** | 185 |
|  |  |  | WoS | **(ALL=((surrogate measures OR proxy measures OR alternate measures))) AND ALL=((skeletal muscle mass)) and Geriatrics Gerontology or Sport Sciences or Gerontology or Rehabilitation (Web of Science Categories)** | 65 |
