## Supplementary material for "Sarcopenia Assessment in Resource-Constrained Settings: Expert Agreement on Surrogate Measures": Online resource 2

### Surrogate measures of Muscle Strength

Thank You for accepting the invitation to participate in our research work.

I am currently pursuing my PhD at the Manipal College of Health Professions, Manipal Academy of Higher Education, India. My research focuses on the **"Design and Development of an mHealth Application for the Detection of Sarcopenia in Older Adults."** Since the current diagnostic criteria for Sarcopenia involve equipment that may not be readily available or affordable at all facilities, our goal is to create a reliable and user-friendly tool that is accessible to everyone. This will ultimately assist in the early detection and management of sarcopenia.

The research is divided into three key phases:

- The first phase involves a comprehensive literature review to identify surrogate markers for muscle strength, muscle mass, and physical performance. This will be followed by content validation by gathering expert opinions to assess their relevance as a surrogate measure and appropriateness for incorporating into our mHealth application.
- The second phase involves validating the finalized measures and incorporating the same into an application.
- The third phase involves evaluating the diagnostic accuracy of the application against the AWGS 2019 standard criteria.

We have completed the literature review and listed below the items that can be incorporated into the application without external hardware. Each surrogate is to be rated on a four-point

1

I hereby give consent *

◯ Yes

◯ No

#### LITERATURE REVIEW

We searched the following five databases: PubMed, Embase, CINAHL, Web of Science, and Scopus, using a set of pre-validated keywords. We also hand-searched articles from bibliographies and used the Google Scholar search engine. The identified records were imported into Rayyan and deduplicated, followed by title and abstract screening. Ultimately, we identified 17 studies relevant to the search and performed data extraction from which we identified 14 surrogate measures for muscle strength. We have listed below eight items that do not require external hardware. The reference and DOI for the same have been mentioned along with the measure. Kindly rate the following identified surrogate measures of hand grip strength in terms of their relevance to muscle strength and appropriateness to be incorporated in the application.

2

#### Five times chair stand test (n=2);

Standard procedure: Participant is seated on a standard chair with arms folded across the chest. They are instructed to repeat 5 sit-to-stand movements and the examiner records the time taken to complete 5 repetitions. Time is recorded in seconds.

#### Ryu et al. 2022 (DOI:https://doi.org/10.1007/s40520-022-02172-2); Belfield et al. 2024 (DOI: https://doi.org/10.1093/ageing/afae090) *

*Ryu et al., 2022*-Objective: To determine whether chair stand test correlates with physical performance (gait speed) or muscle strength (handgrip strength); to investigate the gender differences.
Method: Participants were asked to complete 5 times chair stand test as quickly as possible without using arms. Handgrip strength was evaluated using digital handgrip dynamometer and usual gait speed over 4m was assessed using automatic gait speed meter.
Conclusion: 5 times chair stand test (CST) time had a higher correlation with gait speed ($r = -0.470$) than handgrip strength ($r = -0.309$); As a proxy tool for muscle strength or physical performance, the 5-times chair stand test fits both gait speed and handgrip strength well, but seems to be a better proxy of gait speed than handgrip strength.

*Belfield et al., 2024*- Objective: To determine prevalence of sarcopenia using HGS or chair stand test.
Method: Body composition was assessed using BIA. Prevalence of sarcopenia was assessed with EWGSOP2 criteria. Handgrip strength (HGS) was assessed using Jamar dynamometer. Chair stand test was assessed by recording time taken for participants to rise 5 times from the chair.
Results- Prevalence of probable sarcopenia using CST (31.7%) was higher than HGS (7.1%) in adults with T2DM. Confirmed cases (5.6% vs 1.6%), and severe (1.0% vs 0.3%) sarcopenia.

| | Strongly agree | Agree | Disagree | Strongly disagree |
|---|---|---|---|---|
| Relevant | ◯ | ◯ | ◯ | ◯ |
| Appropriate | ◯ | ◯ | ◯ | ◯ |

3

#### 30 second chair stand test (n=2);

Standard procedure: The participant is instructed to perform as many repetitions of sit-to-stand as possible within 30 seconds.

**Jones et al. 1998 (DOI- https://doi.org/10.1080/02701367.1999.10608028); Nakatani et al. 2002 (DOI- http://dx.doi.org/10.5432/jjpehss.KJ00003390725)** *

*Jones et al. 1998-*

- Objective: 30 second chair stand test was performed to determine its test-retest reliability as a measure of lower body strength and to assess its validity by comparing it to 1RM leg press using Kaiser Leg press machine.
- Method: Community-dwelling older adults were asked to perform two 30-s chair-stand tests and two maximum leg-press tests, each conducted on separate days 2–5 days apart.
- Conclusion: It was concluded that the 30-s chair stand provides a reasonably reliable and valid indicator of lower body strength in generally active, community-dwelling older adults.

*Nakatani et al. 2002-*

- Objective: To determine the validity and test-retest reliability of 30 second chair stand test (CS-30) for evaluating Lower extremity muscle strength against maximum voluntary isometric knee extension.
- Conclusion- CS-30 shows good test-retest reliability, moderately correlates with max. voluntary isometric and is a useful method of evaluating lower extremity strength in field settings in healthy Japanese older adults.

| | Strongly agree | Agree | Disagree | Strongly disagree |
|---|---|---|---|---|
| Relevant | ◯ | ◯ | ◯ | ◯ |
| Appropriate | ◯ | ◯ | ◯ | ◯ |

4

#### 10 times sit to stand (n=1);

Standard procedure: Participant is seated on a standard chair with arms folded across the chest. They are instructed to repeat 10 sit-to-stand movements and the examiner records the time taken to complete 10 repetitions. Time is recorded in seconds.

##### Hardy et al. 2010 (DOI-https://doi.org/10.1007/bf03324942) *

- Objective: To assess whether chair rise performance on comparison to leg extensor power could be solely considered as a measure of leg power.
- Method: The individual was asked to perform a 10 repetition sit to stand test and with standing balance was assessed as longest time the individual can stand on preferred leg with eyes closed. The leg extensor power i.e. LEP (knee extensor) was assessed using Nottingham Power Rig.
- Conclusion: Leg extensor power and standing balance are both related to chair rise time (men> women) and hence, chair rise time should not be solely considered a proxy for leg power in middle aged populations.

| | Strongly agree | Agree | Disagree | Strongly disagree |
|---|---|---|---|---|
| Relevant | ○ | ○ | ○ | ○ |
| Appropriate | ○ | ○ | ○ | ○ |

5

#### Regression equation (n=1);

#### Angst et al. 2010 (DOI- https://doi.org/10.1186/1471-2474-11-94)

*

The equation is as given below.

- Objective: To determine the predictive power of cofactors and to predict population-based normative grip strength.
- Method: Participants were selected using convenience sampling and grip strength was assessed using Jamar dynamometer. An average of 3 readings per person was entered in the database along with covariates of sex, age, body height, body weight, and demands on the hand due to occupational activity (classified into six categories: beyond sedentary, sedentary, light, medium, heavy, very heavy) as set out in the directory of occupational titles. Coding was 0 = f, 1 = m for sex, age in years, height in cm, weight in kg, 0 = beyond sedentary, 1 = sedentary, 2 = light, 3 = medium, 4 = heavy, 5 = very heavy for the variable of occupational demands. Multiple variables were collected along with details of occupational activity using a standardized questionnaire.
- Conclusion: The five easy-to-measure cofactors sex, age, body height, categorized occupational demand on the hand, and body weight provide a highly accurate prediction of normative grip and pinch strength.

$$\text{Grip strength (kg)} = -28.148 + 12.500\ \text{sex} + 0.372\ \text{age} - 0.005\ \text{age}^2 + 0.304\ \text{height} - \text{weight} + 0.001\ \text{weigth}^2 + 12.293\ \text{occupation} - 5.865\ \text{occupation}^2 + 0.897\ \text{occupation}^3$$

| | Strongly agree | Agree | Disagree | Strongly disagree |
|---|---|---|---|---|
| Relevant | ◯ | ◯ | ◯ | ◯ |
| Appropriate | ◯ | ◯ | ◯ | ◯ |

6

#### Blink rate (n=1);

It is calculated as the number of blinks per minute.

##### Bahsi et al. 2021 (DOI- https://doi.org/10.1007/s11845-020-02454-6) *

- Objective: To investigate a relationship between blink rate and grip strength and dynapenia.
- Method: Patients were instructed to blink continuously for 60s and the observer had to note down the count (using digital chronometer) at 15s, 30s, and 60s. Three trials were performed.
- Conclusion-The authors find that in patients where it is not possible to use the hand dynamometer to measure grip strength, the blink rate can be used as an alternative test to detect dynapenia. They also report the cut-off for blink rate at 15 s is ≤40.5 with a sensitivity and specificity of 70.30% and 43.30% respectively.

| | Strongly agree | Agree | Disagree | Strongly disagree |
|---|---|---|---|---|
| Relevant | ◯ | ◯ | ◯ | ◯ |
| Appropriate | ◯ | ◯ | ◯ | ◯ |

7

#### VibPress (n=1);

An application developed to detect the pressure exerted using the accelerometer and vibration sensor in mobile phone.

##### Hwang et. al. 2013 (DOI-https://doi.org/10.1145/2493190.2493193) *

This research states that the amount of pressure on a mobile device can be approximated using accelerometer to measure spatial displacement generated when internal vibration motor vibrates. It furthers states that a software can be built to measure the amount of vibration so that the input pressure can be reliably estimated in the device screen. They have used two different gestures- tap on the screen or Squeeze the sides. When a user touches a button on the capacitive screen, VibPress activates the internal linear vibration motor with a continuous pulse at maximum amplitude which is demonstrated on the screen in different forms.

| | Strongly agree | Agree | Disagree | Strongly disagree |
|---|---|---|---|---|
| Relevant | ◯ | ◯ | ◯ | ◯ |
| Appropriate | ◯ | ◯ | ◯ | ◯ |

8

#### Calf-raise senior test (n=1);

#### Andre H et al. 2016 (DOI- http://dx.doi.org/10.2147/CIA.S115304) *

- Objective: To develop a protocol for field test to assess strength and power of plantar flexors in older adults and to evaluate its validity and reliability.
- Method: Participants were instructed to perform calf-raise movement continuously for 30 seconds in each task selected in random order.
- The tasks are as follows: A) unilateral limb support B) bilateral limb support C) predetermined rate of 60 repetitions/min defined by metronome and D) maximum repetitions in 30 seconds using a self-determined pace. The participants could support their finger on a wall. They were asked to raise their heels as high as possible during the test, maintaining the range of movement by placing their head against an upper bar. For validity and reliability, a battery of functional fitness test, calf-raise senior (CRS) test and strength assessment using Biodex System III isokinetic dynamometer in dominant foot was done.
- A linear regression analysis also shows that the number of repetitions could predict the max PF isometric strength in older participants. The equation is as follows: max isometric strength (MISM) =8.273+1.485× (CRS result).
- Conclusion: The authors find that this test can be a good indicator of ankle strength in older adults.

| | Strongly agree | Agree | Disagree | Strongly disagree |
|---|---|---|---|---|
| Relevant | ◯ | ◯ | ◯ | ◯ |
| Appropriate | ◯ | ◯ | ◯ | ◯ |

9

**Sit-to-stand test (n=1);**

**Ruiz-Cardenas et al. 2023 (DOI- https://doi.org/10.2196/47873); (DOI- https://doi.org/10.1007/s40520-023-02451-6)**

The sit-to-stand app is developed to classify older community-dwelling adult as sarcopenia using the muscle power as a proxy for muscle strength and calf circumference as proxy for muscle mass. Method: The phone is mounted on a tripod at a distance from the participant. They are then asked to perform a sit to stand movement which is recorded in the camera. The video is then analysed to assess the time taken in seconds. A regression equation is used to calculate the muscle power which is then used along with calf circumference to detect sarcopenia.Conclusion: It has a sensitivity of 0.7 to 0.83, specificity of 0.77 to 0.95 and diagnostic accuracy of >76% for women and > 86% for men.

| | Strongly agree | Agree | Disagree | Strongly disagree |
|---|---|---|---|---|
| Relevant | ○ | ○ | ○ | ○ |
| Appropriate | ○ | ○ | ○ | ○ |

10

In addition to the items listed above, we kindly request that you provide any potential surrogate measures for muscle strength that you consider appropriate or relevant, and which could be effectively incorporated into an application. *

**Thank you for your response.** We sincerely appreciate you taking the time to review the study and provide your response. We respect your decision not to participate at this time. If you ever have any questions or would like to learn more about the study in the future, please feel free to reach out. Thank you once again for your time.

Best regards,

Ms. Meghna Prabhu,
Research Scholar,
Department of Physiotherapy,
Manipal College of Health Professions,
Manipal Academy of Higher Education,
Manipal, Udupi.
Karnataka, India.



This content is neither created nor endorsed by Microsoft. The data you submit will be sent to the form owner.

Microsoft Forms

### Surrogate measures for Physical Performance

Thank You for accepting the invitation to participate in our research work.

I am currently pursuing my PhD at the Manipal College of Health Professions, MAHE, India. My research focuses on the **"Design and Development of an mHealth Application for the Detection of Sarcopenia in Older Adults."** Since the current diagnostic criteria for Sarcopenia involve equipment that may not be readily available or affordable at all facilities, our goal is to create a reliable and user-friendly tool that is accessible to everyone. This will ultimately assist in the early detection and management of sarcopenia.

The research is divided into three key phases:

- The first phase involves a comprehensive literature review to identify surrogate markers for muscle strength, muscle mass, and physical performance. This will be followed by content validation by gathering expert opinions to assess their relevance as a surrogate measure and appropriateness for incorporating into our mHealth application.
- The second phase involves validating the finalized measures and incorporating the same into an application.
- The third phase involves evaluating the diagnostic accuracy of the application against the AWGS 2019 standard criteria.

We have completed the literature review and listed below the items that can be incorporated

1. I hereby give consent *

- ◯ Yes
- ◯ No

2. Kindly provide your email address *

[ ]

#### LITERATURE REVIEW

We searched the following five databases: PubMed, Embase, CINAHL, Web of Science, and Scopus, using a set of pre-validated keywords. We also hand-searched articles from bibliographies and used the Google Scholar search engine. The identified records were imported into Rayyan and deduplicated, followed by title and abstract screening. Ultimately, we identified 17 studies relevant to the search and performed data extraction from which we identified 5 surrogate measures for physical performance that do not require external hardware. The reference and DOI for the same have been mentioned along with the measure. Kindly rate the following identified surrogate measures in terms of their relevance to physical performance and appropriateness to be incorporated in the application.

3. **5 times sit-to-stand test (n=4);**

Standard procedure: Participant is seated on a standard chair with arms folded across the chest. They are instructed to repeat 5 sit-to-stand movements and the examiner records the time taken to complete 5 repetitions. Time is recorded in seconds.

**Huang K et al. 2018 (DOI- https://doi.org/10.12968/ijtr.2018.25.4.158);**
**Yee XS et al. 2021 (DOI- https://doi.org/10.1186/s11556-020-00255-5);**
**Nishimura T et al. 2017 (DOI- https://doi.org/10.1111/ggi.12766);Pinheiro P et al. 2016 (DOI-https://doi.org/10.1007/s12603-016-0676-3)** *

*Nishimura T et al. 2017-*

- Objective: To develop a formula to estimate chair stand time based on gait speed and to determine its accuracy to see if it can be a surrogate for gait speed.
- Method: Chair stand time was recorded as time taken to stand from chair 5 times. Gait speed was measured over a 6m distance.

*Pinheiro P et al. 2016 -*

- Objective: To determine the association between chair stand test and sarcopenia and to identify whether it can be a good screening tool for sarcopenia in community-dwelling elderly women.
- Method: The participants were asked to cross their arms across their chests and perform 5 repetitions of sit-to-stand.

*Huang K et al, 2018-*

- Objective: To investigate the reliability and validity of 5 repetition sit-to-stand test (FRSTST) in adult kidney transplant recipients.
- Method: 57 adult patients within 1 year of transplant assessed by 2 testers for FRSTST.

*Yee XS et al. 2021-*

- Objective: To determine the relationship of 5 times sit-to-stand test (5TSTS) and 30 sec Chair Stand Test (30CST) with grip strength and physical performance;
- Method: The participants were instructed to perform 30CST and 5TSTS test.

Conclusion: 5TSTS or CST is a reliable screening tool for sarcopenia, correlates well with physical performance and may be a better surrogate for gait speed rather than handgrip strength.

| | Strongly agree | Agree | Disagree | Strongly disagree |
|---|---|---|---|---|
| Relevant | ◯ | ◯ | ◯ | ◯ |
| Appropriate | ◯ | ◯ | ◯ | ◯ |

4. **Functional Fitness test (n=1);**

**Benton MJ (DOI-https://doi.org/10.1097/phm.0b013e3181aa2ff8)**
*

- Objective: (i) To evaluate the relationship between a battery of four functional fitness field tests and two objective laboratory measures of upper- and lower-body strength in frail, older adults diagnosed with COPD; and (ii) to determine the ability of the field tests to predict absolute strength.
- Method: 40 older adults were asked to perform 2 maximal strength (1RM) tests for upper and lower body. They were also asked to perform Chair stand and Up and Go tests (distance of 8ft).
- Conclusion- Since the chair stand test showed a strong correlation to leg press, it could be considered to be valid surrogate of muscle strength.

| | Strongly agree | Agree | Disagree | Strongly disagree |
|---|---|---|---|---|
| Relevant | ◯ | ◯ | ◯ | ◯ |
| Appropriate | ◯ | ◯ | ◯ | ◯ |

5. **Gait speed (n=1);**
Time taken to walk a specific distance; reported in m/s

**Tiernan C et al. 2024 (DOI- https://doi.org/10.1519/jpt.0000000000000397)** *

- Objective: To determine whether usual or fast gait speed was more strongly associated with physical performance.
- Method: Participants were asked to walk at their usual speed and then to walk as fast and safely as possible. 2 trials were done over a 20m distance with 2 m of acceleration and deceleration zone.
- Conclusion: Strong correlations were observed for fast gait speed compared with usual gait speed with physical performance measures in older adults.

| | Strongly agree | Agree | Disagree | Strongly disagree |
|---|---|---|---|---|
| Relevant | ◯ | ◯ | ◯ | ◯ |
| Appropriate | ◯ | ◯ | ◯ | ◯ |

6. **Anthropometric measures (n=1);**

**Mamphwe P et al 2020 (DOI-https://doi.org/10.1002/ajhb.23324)** *

- Objective: To determine the association between anthropometric measures and physical performance of black adult men and women in South Africa.
- Method: This is a nested cohort study. Anthropometry measures (height, body mass, BMI, calf circumference-CC) were taken at baseline, 5 year and 10 year follow up. Physical Performance (chair stand test, 6 m walk speed and handgrip strength-HGS) was assessed at 10 year follow-up. Basic demographic information (age, sex, education level, and smoking status) was collected.
- Conclusion- BMI and CC in men and women were associated with HGS, but CC was associated with gait speed only in men. They suggest that CC may be a useful predictor of physical performance in black men and to a limited extent in black women.

| | Strongly agree | Agree | Disagree | Strongly disagree |
|---|---|---|---|---|
| Relevant | ◯ | ◯ | ◯ | ◯ |
| Appropriate | ◯ | ◯ | ◯ | ◯ |

7. **eSPPB or qSPPB (n=1);**

Electronic SPPB (eSPPB) or Quick SPPB (qSPPB) is an application that is developed to evaluate physical performance in elderly. eSPPB uses gait, balance and chair stand test while qSPPB uses only chair stand test and gait to assess physical performance.

**Park C et al. 2021 (DOI-https://doi.org/10.3390/s21155147)** *

- Objective: To compare the ability of eSPPB and qSPPB to classify sarcopenia in older adults.
- Methods: Participant was assessed using eSPPB toolkit which consisted of multiple sensors and software developed to assess standing balance, walking speed and chair stand test results. They were classified as sarcopenia using the AWGS 2019 criteria.
- Conclusion: The two-component qSPPB is quicker to administer as compared to the three-component eSPPB and does not significantly alter the classifying ability of the application.

| | Strongly agree | Agree | Disagree | Strongly disagree |
|---|---|---|---|---|
| Relevant | ◯ | ◯ | ◯ | ◯ |
| Appropriate | ◯ | ◯ | ◯ | ◯ |

8. In addition to the items listed above, we kindly request that you provide any potential surrogate measures for physical performance that you consider appropriate or relevant, and which could be effectively incorporated into an application. *

**Thank you for your response.** We sincerely appreciate you taking the time to review the study and provide your response. We respect your decision not to participate at this time. If you ever have any questions or would like to learn more about the study in the future, please feel free to reach out. Thank you once again for your time.

Best regards,

Ms. Meghna Prabhu,
Research Scholar,
Department of Physiotherapy,
Manipal College of Health Professions,
Manipal Academy of Higher Education,
Manipal, Udupi.
Karnataka, India.



This content is neither created nor endorsed by Microsoft. The data you submit will be sent to the form owner.

Microsoft Forms

### Surrogate measures for Muscle Mass

Thank You for accepting the invitation to participate in our research work.

I am currently pursuing my PhD at the Manipal College of Health Professions, MAHE, India. My research focuses on the **"Design and Development of an mHealth Application for the Detection of Sarcopenia in Older Adults."** Since the current diagnostic criteria for Sarcopenia involve equipment that may not be readily available or affordable at all facilities, our goal is to create a reliable and user-friendly tool that is accessible to everyone. This will ultimately assist in the early detection and management of sarcopenia.

The research is divided into three key phases:

- The first phase involves a comprehensive literature review to identify surrogate markers for muscle strength, muscle mass, and physical performance. This will be followed by content validation by gathering expert opinions to assess their relevance as a surrogate measure and appropriateness for incorporating into our mHealth application.
- The second phase involves validating the finalized measures and incorporating the same into an application.
- The third phase involves evaluating the diagnostic accuracy of the application against the AWGS 2019 standard criteria.

We have completed the literature review and listed below the items that can be incorporated into the application without external hardware. Each surrogate is to be rated on a four-point Likert scale. The option for neutral has not been provided to avoid ambiguity and to keep

1. I hereby give consent *

◯ Yes

◯ No

2. Kindly provide your email address *

#### LITERATURE REVIEW

We searched the following five databases: PubMed, Embase, CINAHL, Web of Science, and Scopus, using a set of pre-validated keywords. We also hand-searched articles from bibliographies and used the Google Scholar search engine. The identified records were imported into Rayyan and deduplicated, followed by title and abstract screening. Ultimately, we identified 14 studies relevant to the search and performed data extraction. The most commonly identified measure is calf circumference. The reference and DOI for the same have been mentioned along with the measure. Kindly rate the following identified surrogate measures of muscle mass in terms of their relevance and appropriateness to be incorporated in the application.

3. **Calf circumference (n=1);**
**Kawakami R et al. 2014 (DOI- https://doi.org/10.1111/ggi.12377)**
*

*Kawakami R et al. 2014-*

- Objective: To examine the relationship between calf circumference and DXA-measured muscle mass and to determine its suitability as a surrogate measure for muscle mass to diagnose sarcopenia in middle-aged and older Japanese.
- Method: Measures of height, BMI, Calf Circumference (CC) at maximal girth were taken for two trials on each side.
- Conclusion: CC was positively correlated with Appendicular skeletal Muscle Mass (ASMM) and Skeletal Muscle Index (SMI). It could be a potential surrogate for muscle mass for diagnosing sarcopenia. The suggested cut-off for CC to predict low muscle mass is<34 cm in men and <33 cm in women.

| | Strongly agree | Agree | Disagree | Strongly disagree |
|---|---|---|---|---|
| Relevant | ○ | ○ | ○ | ○ |
| Appropriate | ○ | ○ | ○ | ○ |

4. **Calf Circumference (n=1);**
**Sunyoung K et al. 2018 (DOI- [https://doi.org/10.3346/jkms.2018.33.e151](https://doi.org/10.3346/jkms.2018.33.e151))** *

*Sunyoung K et al. 2018-*

- Objective: To identify the optimal cut-off value of calf circumference as a surrogate for muscle mass and sarcopenia in Korean older adults.
- Method: CC was measured in standing position at maximal girth.
- Conclusion: ASMI and CC showed positive correlation. Optimal value for cut-off was 32cm (males, sensitivity 75%, specificity 83%; females, sensitivity 85%, specificity 57%).

| | Strongly agree | Agree | Disagree | Strongly disagree |
|---|---|---|---|---|
| Relevant | ◯ | ◯ | ◯ | ◯ |
| Appropriate | ◯ | ◯ | ◯ | ◯ |

5. **Calf circumference (n=2);**

**Kiss CM et al 2024 (DOI- https://doi.org/10.1007/s40520-024-02694-x);**
**Ukegbu PO et al. 2018 (DOI- https://doi.org/10.1080/16089677.2018.1518825)** *

*Kiss CM et al. 2024-*

- Objective: To determine the correlation between CC and ASMI among geriatric inpatients.
- Method: CC was assessed in sitting and supine position on the right side at maximal calf girth. Three readings were recorded.
- Conclusion: CC and ASMI were found to be positively correlated. Hence, CC can be a dependable screening tool if imaging is not available.

*Ukegbu PO et al. 2018-*

- Objective: To investigate the relationship between CC and ASMI, and to determine whether CC could be used to diagnose sarcopenia in older black South African women.
- Method: CC was recorded in standing position at maximal girth. Handgrip strength was assessed using Jamar hand dynamometer and gait speed with 6m test.
- Conclusion: Positive correlation was observed between CC and ASMI ($r = 0.84$; $p < 0.001$). CC is a useful surrogate of ASM demonstrating good sensitivity and moderate specificity to detect low muscle strength in older black South African women.

| | Strongly agree | Agree | Disagree | Strongly disagree |
|---|---|---|---|---|
| Relevant | ◯ | ◯ | ◯ | ◯ |
| Appropriate | ◯ | ◯ | ◯ | ◯ |

6. **Calf circumference and regression equation (n=1);**

**Hwang AC et al. 2018 (DOI- https://doi.org/10.1016/j.jamda.2017.11.016)** *

- This report is a secondary cross-sectional data analysis from a longitudinal aging study, comprising of 1839 community-dwelling older adults. Their objective was to determine whether calf circumference can provide an accurate estimation of Appendicular Skeletal Muscle Mass (ASM).
- Method: Calf circumference was measured at maximal girth on both sides while ASM was measured using DXA. The prediction equation was ASM (kg) = [-0.028*age (year)]+ [-3.973*sex (men = 1,women = 2)] + (0.097*weight) + [0.148*height (cm)] + 0.147*calf circumference (cm)-8.734.
- Conclusion: Strong correlation between calf circumference and ASM among middle aged and older adults in Taiwan.

| | Strongly agree | Agree | Disagree | Strongly disagree |
|---|---|---|---|---|
| Relevant | ◯ | ◯ | ◯ | ◯ |
| Appropriate | ◯ | ◯ | ◯ | ◯ |

7. In addition to the items listed above, we kindly request that you provide any potential surrogate measures for physical performance that you consider appropriate or relevant, and which could be effectively incorporated into an application. *

### Thank you for your response.

We sincerely appreciate you taking the time to review the study and provide your response. We respect your decision not to participate at this time. If you ever have any questions or would like to learn more about the study in the future, please feel free to reach out. Thank you once again for your time.

Best regards,

Ms. Meghna Prabhu,
Research Scholar,
Department of Physiotherapy,
Manipal College of Health Professions,
Manipal Academy of Higher Education,
Manipal, Udupi.
Karnataka, India.



This content is neither created nor endorsed by Microsoft. The data you submit will be sent to the form owner.

Microsoft Forms
