## Supplementary material for "Sarcopenia Assessment in Resource-Constrained Settings: Expert Agreement on Surrogate Measures": Online resource 3

**Journal: PLOS Digital Health**

**Authors: Ms. Meghna Suresh Prabhu, Dr. Sucheta V Kolekar, Olivier Bruyere, Reshma A Merchant, Sanjay Kalra, Shweta Gore, Nimit Agarwal, Dr Girish N***

***Dr. Girish N (PhD) (corresponding author),**

Additional Professor,

Department of Physiotherapy, Manipal College of Health Professions,

Manipal Academy of Higher Education,

Karnataka, India.

ORCID: 0000-0003-2181-5332

Contact details: +91 9886782114

**Online Resource 2**

Measures identified for muscle strength

| Measure | Author, Year, Country | Age, Gender,  Study settings,  Sample size | Test Measure | Surrogate measure | Findings |
| --- | --- | --- | --- | --- | --- |
| Five times sit-to-stand (5STS) | Ryu et al. 2022, Korea | 70-84 years, Both,  community-dwelling,  N=1416 | HGS using  digital handgrip dynamometer;  Usual gait speed over 4m with 1.5m of acceleration & deceleration | 5CST as quickly as possible without using arms. Classified as failed if they cannot complete. | The 5STS could be a proxy for both gait speed and handgrip strength, but it seems to be a better proxy of gait speed than handgrip strength. |
|  | Belfield et al. 2024, United Kingdom | 18-75 years, Both,  community, N= 732 | HGS, Body composition, and 5STS | HGS, 5STS | Sarcopenia prevalence via 5STS is higher than with HGS in T2DM, likely due to adiposity. |
| 30-second chair stand test (30s CST) | Jones et al. 1998, United States | Mean age = 70.5 years, Both,  Community, N=76 | Kaiser Leg Press | 30-sec chair stand test | The 30s CST has good test-retest reliability and also gives a reliable and valid indication of lower body strength in generally active, community-dwelling older adults. |
|  | Nakatani et al. 2002, Japan | 60-87 years, Both,  community, N=486 | Maximum voluntary isometric knee extension | 30-second chair stand test | 30s CST has a high test-retest reliability correlation and is useful to assess lower extremity muscle strength in Japanese elderly men and women in a field setting. |
| 10STS | Hardy et al. 2010, UK | 53 yrs,  Both,  Community-dwelling, N= 174 | Knee extensor Power using Nottingham Power Rig (NPR) | Standing balance, chair rising performance (10STS) | LEP and standing balance are both related to chair rise time in men, suggesting that chair rise time should not be thought purely as a proxy for leg power in middle-aged populations |
| Regression equation | Angst et al. 2010, Switzerland | 18-96 years,  Both,  community-dwelling,  N= 978 | Grip strength- Jamar dynamometer | Predictive equation using co-factors such as height, weight, sex, age, and occupational demand on hand | The five cofactors provided a highly accurate prediction of normative grip strength. |
| Blink rate | Bahsi et al. 2021,  Turkey | >65 years, Both,  Outpatient clinic,  N=355 | HGS; GRIP-D, grip strength dynamometer; 4m gait speed test | Blink rate; blinking counts of 15s, 30s, and 60s were noted by observer using digital chronometer | This study shows that blink rate can be used as an alternative test in patients with limited mobility and where HGS testing is not possible. |
| VibPress | Hwang et al. 2013,  South Korea | 26-33 years,  Both,  Feasibility study,  N=4 | NS | Squeeze and press gesture on the phone | VibPress demonstrates multiple pressure levels, which may provide an estimate of the different pressure levels generated by a press or squeeze gesture. No clinical studies are reported to determine its reliability and validity. |
| Calf-raise senior test (CRS) | Andre et al. 2016,  Portugal | >65 years, Both,  community-dwelling,  N= 41 | Isokinetic and isometric plantarflexion strength in the dominant foot using Biodex System III | Calf-raise senior test;  Linear regression analysis using the equation MISM=8.273 + 1.485 x (CRS result) | Calf-raise senior test can be a good indicator of ankle strength in older adults in field settings. |
| Sit-to-stand app | Ruiz-C et al. 2023,  Spain | >60 years,  Both,  Community-dwelling,  N= 686 | Standard criteria of HGS | Sit-to-stand movement recorded by camera;  Muscle power calculated by the app | The app showed good diagnostic performance for detecting sarcopenia in well-functioning Spanish community-dwelling older adults. |

NA- Not Applicable; NS- Not Specified; T2DM- Type 2 Diabetes Mellitus; LEP- Lower Extremity Power; CST- chair stand test; HGS- Handgrip strength; MISM- Maximal isometric
