## Supplementary material for "Sarcopenia Assessment in Resource-Constrained Settings: Expert Agreement on Surrogate Measures": Online resource 4

**Journal: PLOS Digital Health**

**Authors: Ms. Meghna Suresh Prabhu, Dr. Sucheta V Kolekar, Olivier Bruyere, Reshma A Merchant, Sanjay Kalra, Shweta Gore, Nimit Agarwal, Dr Girish N***

***Dr. Girish N (PhD) (corresponding author),**

Additional Professor,

Department of Physiotherapy, Manipal College of Health Professions,

Manipal Academy of Higher Education,

Karnataka, India.

ORCID: 0000-0003-2181-5332

Contact details: +91 9886782114

**Online Resource 3**

Measures identified for physical performance

| Measure | Author, Year, Country | Age, Gender | Study Design, settings, Sample size | Test Measure | Surrogate measure | Findings |
| --- | --- | --- | --- | --- | --- | --- |
| Five times sit-to-stand (5STS) | Huang K et al. 2018;  Singapore | Between 19-69 years;  Both | Cross-sectional, Hospital  N=56 | Quadriceps muscle strength with digital myometer,  Physical function (STS60; DASI) | 5STS | In kidney transplant patients, the 5STS test proves to be a good measure of physical function, but not lower limb muscle strength. |
|  | Yee XS et al. 2021, Singapore | >50 years,  Both | Cross-sectional, community-dwelling;  N=887 | SPPB, Habitual gait speed, 6MWT, TUG, HGS | 5STS and 30 sec chair stand test (30s CST) | The sit-to-stand tests give a better representation of physical performance in older adults as compared to muscle strength. |
|  | Nishimura et al. 2016, Japan | >40 years, Both | Cross-sectional, community-dwelling;  N=629 | Gait speed | 5STS;  estimated formula: 5CST=-8.41 * gait speed + 20.0 (R^2^=0.34) | CST can be a useful surrogate of gait speed as it is feasible for small spaces, and hence may assist in sarcopenia screening. |
|  | Pinheiro et al. 2016, Brazil | ≥ 60 years, females | Cross-sectional, community-dwelling;  N= 173 | 2.44m gait speed | Chair stand test (CST) | This study indicates that it could be a possible screening tool for sarcopenia. |
| Functional fitness test | Benton et al. 2009, US | 70 years, Both | Cross-sectional; N=40 | 1RM for upper limb strength (incline chest press), 1RM for lower limb strength (leg press), and | Functional fitness testing: arm curl, chair stand, (max reps in 30s), and Up & Go | The arm curl test and the chair stand test may be used as measures for the strength of the upper and lower body, respectively, in the absence of objective laboratory tests. |
| Gait speed | Tiernan et al. 2024,  US | 60-87 years, Both | Cross-sectional, Community-dwelling,  N=57 | step execution time, 6MWT, knee extension strength, ABC-6 | usual gait speed, and fast gait speed | Fast gait speed showed better correlation with measures of physical performance as compared to the usual gait speed and could be considered as a key predictor of physical performance measures. |
| Anthropometric measures | Mamphwe et al. 2019, South Africa | 32-93 years, Both | Nested Cohort, Community-dwelling;  N= 774 | chair stand, 6 m walk speed, and handgrip strength (HGS) | height, Body Mass Index (BMI), Calf circumference (CC) on right calf at max girth | CC predicted walking speed in men. CC may be a useful tool for assessing physical performance, especially in Black men. |
| eSPPB or eQPPB | Park et. al, 2021, Korea | Median age = 78 years,  Both | Cross-sectional,  Community-dwelling,  N=124 | Gait speed | eSPPB-to measure balance, 5STS and 4m walk test;  eQPPB- to assess 5STS and 4m walk test | Omitting balance tests may reduce test time without significantly affecting the eSPPB's ability to classify sarcopenia. |

STS- Sit-to-stand test; DASI- Duke Activity Status Index; RM- repetition maximum; 6MWT: 6-minute walk test; ABC-6- Activities specific balance confidence scale; TUG- Timed Up & Go test; eSPPB- electronic Short Physical Performance Battery; eQPPB- electronic Quick Physical Performance Battery
