## Supplementary material for "Sarcopenia Assessment in Resource-Constrained Settings: Expert Agreement on Surrogate Measures": Online resource 5

**Journal: PLOS Digital Health**

**Authors: Ms. Meghna Suresh Prabhu, Dr. Sucheta V Kolekar, Olivier Bruyere, Reshma A Merchant, Sanjay Kalra, Shweta Gore, Nimit Agarwal, Dr Girish N***

***Dr. Girish N (PhD) (corresponding author),**

Additional Professor,

Department of Physiotherapy, Manipal College of Health Professions,

Manipal Academy of Higher Education,

Karnataka, India.

ORCID: 0000-0003-2181-5332

Contact details: +91 9886782114

**Online Resource 4**

Measures identified for muscle mass

| Measure | Author, Year, Country | Age, Gender,  Study settings, Sample size | Test Measure | Surrogate measure | Findings |
| --- | --- | --- | --- | --- | --- |
| Calf circumference (CC) | Kawakami et al, 2014, Japan | 40-89 years, Both,  Community-dwelling,  N= 526 | Skeletal Muscle Index (SMI) using DEXA scanner | Calf circumference-  2 trials in the standing position along the maximal girth of the calf | CC was positively correlated with ASMM and SMI, and could be used as a surrogate marker of muscle mass for diagnosing sarcopenia. |
|  | Sunyoung et al., 2018, Korea | 70-80 years,  Both,  Community-dwelling,  N=657 | Appendicular skeletal mass (ASM) and fat mass of all four limbs | upper arm circumference, CC, WC, | CC may be a good indicator of sarcopenia in the Korean elderly. |
|  | Kiss et al, 2024,  Switzerland | Mean age =83.5;  Both,  Hospital, | BIA | Calf circumference | This study demonstrates that CC and ASMI are positively correlated and also establishes gender-specific cut points. |
|  | Ukegbu et al., 2018,  South Africa (SA) | >45 years,  Females,  Community-dwelling,  N=247 | ASM < 4.94 kg/m2 (cut-off for SA population) with DEXA | Calf Circumference | CC of 30 cm is proposed as a simple and inexpensive way to predict, screen or diagnose sarcopenia in black women in low-resource health settings. This could facilitate timely intervention and prevention. |
| Regression equation | Hwang et al, 2018,  Taiwan | >50 years,  Both,  Community-dwelling,  N=1839 | DEXA | ASM (kg) = [-0.028*age (year)] + [-3.973*sex (men = 1, women = 2)] + (0.097*weight) + [0.148*height (cm)] + 0.147*calf circumference (cm)-8.734. | CC could be an easy-to-use screening tool for ASM in the community and a simple instrument to promote the awareness of sarcopenia. |
| Anthropometric measures | Sousa-Santos et al. 2021,  Spain | >65 years, Both, Community-dwelling,  N= 159 | ASM and SMM | height, weight, MAC, WC, CC, triceps skinfold thickness, BMI, MAMC = (MAC − [3.14 × TSF] | BIA poses a suitable alternative to DEXA for muscle mass assessment in sarcopenia diagnosis. CC also serves as a reliable indicator for identifying sarcopenia. |
|  | Ling et al, 2021,  Australia | Both,  Cohort, Community-dwelling, N=572 | SMM (random sample of 78) | height, weight, body circumferences (MUAC, WC, HC, CC), BMI, W/H ratio | This study shows that simple anthropometric measures such as mid-upper arm and CC can be a proxy for body composition in a geriatric outpatient setting. |

NA- Not Applicable; NS- Not Specified; ASM- appendicular skeletal mass; SMM- skeletal muscle mass; DEXA- dual energy x-ray absorptiometry; BIA-Bioelectrical Impedance Analysis; WC- Waist circumference; HC- hip circumference; MAC- mid-arm circumference; TSF- triceps skin fold; BMI- body mass index; MUAC- mid upper-arm circumference; WHtR- waist to height ratio
