## Supplementary material for "Sarcopenia Assessment in Resource-Constrained Settings: Expert Agreement on Surrogate Measures": Online resource 6

**Title: Identification and Expert Agreement on Surrogate Measures for Sarcopenia in Older Adults in Low-Resource Settings**

**Journal: European Geriatric Medicine**

**Authors: Ms. Meghna Suresh Prabhu, Dr. Sucheta V Kolekar, Olivier Bruyere, Reshma A Merchant, Sanjay Kalra, Shweta Gore, Nimit Agarwal, Dr Girish N***

***Dr. Girish N (PhD) (corresponding author),**

Additional Professor,

Department of Physiotherapy, Manipal College of Health Professions,

Manipal Academy of Higher Education,

Karnataka, India.

ORCID: 0000-0003-2181-5332

Contact details: +91 9886782114

**Online Resource 5**

Details of Experts:

| Sr. No | Affiliation | Specialization | Country | LMIC/HIC | h-index (Scopus) |
| --- | --- | --- | --- | --- | --- |
| 1 | Professor, Research Unit in Public Health, Epidemiology and Health Economics, University of Liege, Belgium | Clinical Trials  Epidemiology  Geriatrics | Belgium | HIC | 88 |
| 2 | Associate Professor, Division of Geriatric Medicine, National University Hospital, Singapore | Geriatrics  Internal Medicine (General Medicine) | Singapore | HIC | 33 |
| 3 | Treasurer, International Society of Endocrinology (ISE)  Vice President, South Asian Obesity Forum (SOF).  Consultant, Department of Endocrinology, Bharti Hospital, Karnal, India | Endocrine disorders  Glucose  Insulin, Sarcopenic obesity | India | LMIC | 55 |
| 4 | Chair of the Geriatric Department at the Hospital Universitario Ramón y Cajal, Madrid, Spain.  Associate Professor of Geriatrics at the Universidad Europea de Madrid, Madrid, Spain. | Geriatrics, Medicine, Sarcopenia, Frailty | Spain | HIC | 72 |
| 5 | Public Health Aging Research & Epidemiology (PHARE) Group  Research Unit in Clinical Pharmacology and Toxicology (URPC)  NAmur Research Institute for LIfe Sciences (NARILIS)  Department of Biomedical Sciences- Faculty of Medicine, University of Namur, Belgium | M. Sc, M. PH, PhD, sarcopenia, osteoporosis, patients’ preferences, public health, measurement properties. | Belgium | HIC | 53 |
| 6 | Research Fellow, Geriatrician, The University of Melbourne, The Royal Melbourne Hospital, Australia | MBBS, MPHTM PhD, Epidemiology  Geriatrics | Australia | HIC | 17 |
| 7 | Associate Professor, Department of Physical Therapy,  MGH Institute of Health Professions, Boston | Certified Clinical Specialist in Geriatric Physical Therapy (GCS) from the American Board of Physical Therapy Specialties (ABPTS). | United States of America | HIC | 7 |
| 8 | Clinical Associate Professor of Medicine  Geriatrics, Internal Medicine and Hospital Medicine.  University of Arizona-College of Medicine-Phoenix,  Banner University Medical Center-Phoenix  Eller College of Management. | Geriatrics | United States of America | HIC | 4 |
| **Total invited: 34; Accepted: 9; Completed: 8; Countries: 6; Experience (years): >10; h index (mean): 41.12** | | | | | |

LMIC- Low and middle-income countries; HIC- High-income countries
